# An agent-based model of spread of a pandemic with validation using COVID-19 data from New York State

**DOI:** 10.1101/2020.10.19.20215517

**Authors:** Amitava Datta, Peter Winkelstein, Surajit Sen

## Abstract

We introduce a simple agent based model where each agent carries an effective viral load that captures the instantaneous state of infection of the agent and simulate the spread of a pandemic and subsequently validate it by using publicly available COVID-19 data. Our simulation tracks the temporal evolution of a virtual city or community of agents in terms of contracting infection, recovering asymptomatically, or getting hospitalized. The virtual community is divided into family groups with 2-6 individuals in each group. Agents interact with other agents in virtual public places like at grocery stores, on public transportation and in offices. We initially seed the virtual community with a very small number of infected individuals and then monitor the disease spread and hospitalization over a period of fifty days, which is a typical time-frame for the initial spread of a pandemic. An uninfected or asymptomatic agent is randomly selected from a random family group in each simulation step for visiting a random public space. An uninfected agent contracts infection if the public place is occupied by other infected agents. We have calibrated our simulation rounds according to the size of the population of the virtual community for simulating realistic exposure of agents to a contagion. Our simulation results are consistent with the publicly available hospitalization and ICU patient data from different communities of varying sizes in New York state. Our model can predict the trend in epidemic spread and hospitalization from a set of simple parameters and could be potentially useful in exploring strategies to keep a community safe.

## Introduction

Epidemiological models for spread of infectious diseases are very important for taking policy decisions for mitigating the spread of an outbreak of a highly infectious disease. Perhaps the earliest work on such modelling was by Kermack and McKendrik (1). They introduced a class of models referred to as compartmental models (2). Nowadays these models are often referred to as the SIR (Susceptible, Infected, Recovered) models or some equivalent thereof (3). In these models the entire population is divided into three compartments according to these labels and the transitions of people from one compartment to another determine the rates of infected and recovered people. Infected people may infect susceptible people, and infected people may either develop immunity, or may be removed from the population due to death. Simi-larly new individuals enter the population due to birth. However birth and death may be ignored and the population may be considered as conserved during rapid spread of a disease without incurring significant calculational errors. The transition rates between the compartments are the free parameters in this model.

The SIR model has been extended in several different ways. For example only two compartments, susceptible and infectious are used when individuals do not develop immunity in case of diseases like influenza. The SIRD (susceptible-infectious-recovered-deceased) model differentiates between the recovered and deceased compartments. Infected individuals can either recover or die. The transitions over time are modeled as first order differential equations in all SIR models. These models also have been discretized and simulated (4).

In spite of their success, the compartmental models are simplistic and consider only homogeneous population groups within each compartment. As a result, complex interactions between individuals or complex behavior of individuals are hard to capture in these models (5). The spread of a pandemic is a complex phenomenon that is determined largely by the interactions between individuals in a community and for example the way a virus can remain active on the surfaces of inanimate objects. Compartmental models can typically capture a global effect that is determined by the transition rates between the compartments, without any finer control at the individual level. For example, one of the most important public policies often invoked during a pandemic outbreak is typically some form of social distancing. Moreover these measures must be optimized to prevent disrupting economic activities as well as to reduce the spread as much as possible. It is difficult to incorporate such social distancing measures in the compartmental models. The only possibility is to introduce some global parameters retrospectively assuming the model fits the data perfectly. In other words, these models are not naturally best suited to make forward predictions. Furthermore, it is unrealistic to assume that each member in a population will behave in an effectively uniform (rationally or irrationally) homogeneous way in a pandemic situation. It is difficult to capture these individual variations in behavior through global parameters, even though such individual behaviors may have global effects in the spread of a pandemic. These shortcomings of the compartmental models were pointed out by Epstein (6) and Parker and Epstein (7). They argued that the assumption of a homogeneous nature of population is in particular unrealistic in compartmental models, since complexity of interactions among individuals through their access to public places and social networks are quite often difficult to capture in these models. On the other hand agent-based modeling can capture these nuances of interactions in a society more accurately. National Institute of Health’s Models of Infectious Disease Agent Study (MIDAS) program (8) based on agent based simulation helped in formulating policies during the avian flu or H5N1 pandemic.

Parker and Epstein (7) reported a global scale agent-based simulation of six billion people for modeling the spread of a pandemic. While the spread of a pandemic is not restricted by national borders in today’s global world, quite often the public policies for preventing the spread of a pandemic are dictated by local situations within a country, state or even a city. Hence it is an important and interesting question to examine whether agent-based models can simulate local situations accurately (9, 10). In particular, an accurate simulation based on a local trend of the spread of a pandemic can give the authorities the scope to frame local policies regarding social distancing measures to curb the future spread of a pandemic. Local and national governments are grappling with this problem of framing appropriate national and local policies rapidly in many countries around the world, including the USA, India, China and the countries in the European Union.

Validating a model of pandemic spread is a challenging problem. It is very difficult to estimate the extent of the spread, as many infected individuals remain asymptomatic. Testing is often reactive, rather than proactive. Individuals are mostly tested when they display symptoms, hence positive cases identified through testing is often biased. One possibility is to consider death rates due to a pandemic, as has been done in many works such as in Ref. (11). However the death rate alone may not be a reliable measure of the spread of a pandemic, due to the fact that quite often older individuals with other health problems (so called co-morbidity) succumb to a new infection disproportionately. Further, it is likely that the number of COVID deaths may be under-reported and there can be significant lags between the actual time of death and the time the death certificate is issued (12, 13). Hence, validating a model based only on death rate can be problematic and does not capture the spread of infection and in particular the severity of the infected population accurately. The challenging aspect of framing public policy is to capture the severity of infection, as severity puts pressure on the health infrastructure in terms of provisioning hospital beds and intensive care units (ICU). Hence in this paper we consider hospitalization and ICU data for validating our model. As we shall see, our study projects proportions of deaths which seem consistent with those reported in various sources. It is possible to use robust and publicly available data as hospitalization and ICU figures are available from the government health departments, for example, from the New York state government (15). Of course the assumption here is that all severely infected patients can be accommodated in hospitals, which is the case in most developed countries, e.g., (15), and at least in the initial stages of a pandemic in developing countries. It should, however, be noted that in New York as time progressed fewer patients were transferred to the ICUs due to an improved ability to treat these patients in the hospitals. Thus, the proportions of people who ended up in the ICUs did not remain a constant in time (16). However, in our work we have regarded this number as a constant for the sake of simplicity. Further refinements of the model or rendering the proportion of people who are transferred to ICU can indeed be easily varied in our study.

We present an agent-based modeling framework for local communities and cities in this paper. To our knowledge, this work is distinct from other attempts using an agent based approach (see for example in Ref. (17)) We introduce two types of entities in this model: home groups and public places. Each home group consists of 2 − 6 members and each public place has a fixed capacity. We start the simulation with a small percentage of infected agents among the members of the home groups (typically about 0.1% of the total population). In each time step of the simulation, a random member of a randomly chosen home group visits a random public place if all the public places are not filled to capacity. An agent picks up infection from a public place in the presence of other infected agents.

There are over fifty models currently deployed in Covid-19ForecastHub (14) for forecasting the incidence of infection and death due to Covid-19. Most of these models are different adaptations of the SIR model, regression models or ensemble models. The main aim of the models is to accurately predict the spread of infection and deaths by using the data known until now. Hence the objectives of these models are quite different from the model we present in this paper. Our aim is to choose our parameters from the very early trends of the spread and validate the model through precise data on hospitalization and admission to ICU. We believe that the data on the spreading of infection may not be very accurate due to biases in testing. The paper by Keskinocak et al. (18) discusses an agent-based model. However, they mainly use their model to study the effects of social distancing measures and compare their simulation results with data on spreading of infection.

The rest of the paper is organized as follows. We discuss the details of our model and the methodology in section 2, we discuss the results in section 3, and we conclude with discussions and future work in section 4.

### Details of the model

Depending upon the effective viral loads carried, we assume that there are four categories of agents in our simulations. We also let the entire population to be susceptible to infection and allow only a part of the population to have better immune response. This segment of the population can recover from the infection asymptomatically. The other part develops complications.

The effective viral load of category 1 agents reaches a low threshold, stabilizes for a certain time and then a typical agent recovers with a presumed exponential decay of the viral load. The exponential decay is assumed because we chose to assign the gradual reduction of the effective viral load to have a characteristic time scale as opposed to being scale-free. We do not consider immunity in this work, as it is not well established whether there is even any short-term immunity from COVID-19. Hence agents who have recovered from a mild infection may get infected again. The effective viral load of a category 2 agent continues to grow until it reaches a higher value that requires hospitalization. A category 2 agent has two possible outcomes. One possibility is that a category 2 agent may stay in the hospital for a certain number of days and then recovers through an exponential decay of its viral load. Otherwise a category 2 agent is shifted to ICU after a certain period, and then may either transform into a category 3 agent and dies, or recovers from ICU as a category 4 agent, again through an exponential decay of its viral load. Since our simulation time frame is rather limited, we assume that agents in categories 2, 3 and 4 do not visit any public places. We have chosen these categories in order to validate our model in a robust manner given that the data for hospitalization and admission to ICU are quite robust and publicly available (15). Note that we have avoided using the death rate as a quantity that is being tracked in our model except to assume that the number of deaths of the agents in the ICUs should be consistent with the overall number of deaths in the entire period of our simulations. The main result of this paper is that simple agent-based models give accurate results for hospitalization and admission to ICUs based upon parameters that can be constructed through trial and error as well as on directly available data as explained in detail below. Moreover, once the needed parameters are extracted for a given region and the population involved, our model can predict the future trends in hospitalization and admittance to ICU. Thus, our study may allow for the possibility of preparing for future demands on health care, or better provisioning of services.

There is a population of *N* agents divided into *G* home groups, each with 2 − 6 members. We assume that the number of households with a single member is small enough to not have a significant effect on our calculations, a condition that can be relaxed when considering studies on smaller pop-ulations. The size of a home group *g*_*i*_, denoted by |*g*_*i*_| is chosen randomly within this range, although the population *N* is fixed. In other words, *G* is chosen so that Σ|*g*_*i*_| = *N*. The number of time steps *T* in our simulation is based on the notion of a *virtual day*. The simulation is run for 50 days, each day consisting of a number of time steps depending on *N*. This is done to create a realistic situation that at least one member from each home group has a chance to visit a public place like a grocery store for getting provisions at least four times during the simulation period. (However it is not guaranteed that members of each home group will visit public places at least four times due to the stochasticity in the choice of agents in each simulation step). Hence the total number of simulation steps is Σ{|*g*_*i*_|*×* 4} and a day, denoted by *D*, consists of 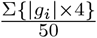 iterations. There are *P* public places *p*_1_, *p*_2_, …, *p*_*P*_, each with capacity *C*. We have kept the capacities of the public places homogeneous with a realistic assumption that an agent comes into contact with only a small number of other agents irrespective of the total capacity of a public place. We vary the public place capacities for implementing social distancing measures in our model. In our realization, an overall lower capacity in public places would expose a smaller number of agents to infection, which is the aim of social distancing.

The viral load profiles of random individuals from various effective viral load categories, those who get mildly sick, those who get sick enough to be hospitalized, then among them those who transition to ICUs and may or may not survive, are reported in Fig 1. Fig. 1 shows the representative agents from each category. The plateau regions of infections in Fig. 1 are between 3 − 6 days, according to our notion of a virtual day, randomly chosen for each agent. The viral load of a category 1 agent increases for one day, that of category 2, 3 and 4 agents increases for three days. The separation between category 3 and 4 agents occurs after one day in the ICU, i.e., category 4 agents lose their viral load through an exponential decay and category 3 agents progress towards death through an exponential rise in viral load.

**Fig. 1.**
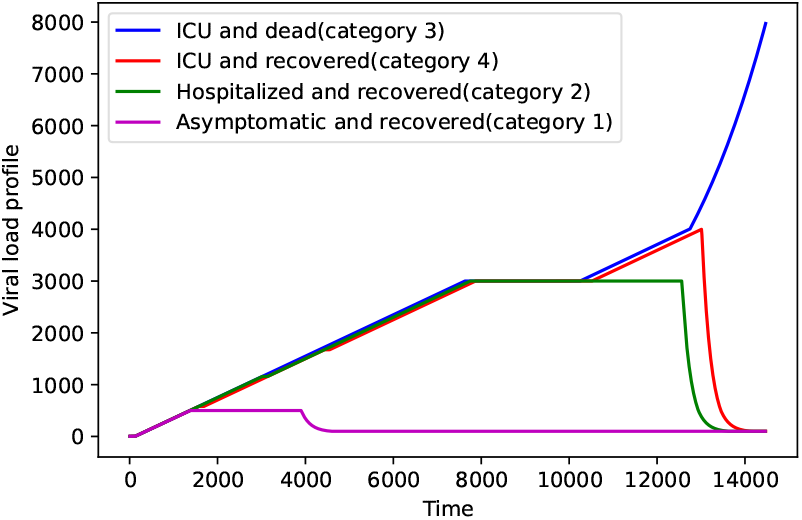
Four categories of agents in our model (see discussion in text).

The life-cycle of an agent is as follows. An agent either stays at home, or visits a public place. The viral load of an uninfected agent is zero and continues to be so when it stays at home. A random agent is selected from a random home group in each simulation step, provided the agent is not a category 2, 3 or 4 agent. Category 1 agents may be chosen at any stage and we assume that these agents are asymptomatic and able to visit public places. Once an agent is chosen, it visits a public place depending on the availability of space in the public place. We randomly choose a public place to accommodate the agent. A simulation step is considered as a *failure* if all the public places are already filled. Once an agent enters a public place, it stays there for a small amount of time, arbitrarily fixed at about 10 minutes according to our calibration. An uninfected agent gets infected if the public place was previously visited by other infected agents, or if there are other infected agents currently visiting the public place. We assume this initial infection to be a small amount of viral load, as a particular agent may visit a public place at most a few times and the duration of the stay at a public place is also short. Finer models of initial infection, e.g., an exponential increase in viral load depending upon how many other infected agents are simultaneously present in the public place, did not show any qualitative differences in our simulation.

Agents spend most of the simulation time in their home groups. An agent enters its home group after returning from a public place and remains there unless chosen again to visit a public place. As we have mentioned before, only uninfected or category 1 agents can visit a public place. While in a home group, the infection level of each agent increases by a unit amount in each time step until a threshold is reached as discussed below. This is the rationale behind our calibration of the simulation time in terms of days, as the viral loads for the transitions in the model are reached due to this increase over specific number of days of simulation time. Hence there are basically two simple transitions in an agent’s life, home to public place and back. We have assumed that the other members of a home group are not infected due to the presence of an infected member, in other words isolation at home is perfect. However it is easy to lift this restriction in our simulation, though the only qualitative effect is an increased rate of infection. Another way of implementing this infection in home groups is to randomly infect a small number of uninfected agents if there is any infected agents in their home groups. However, this also does not produce qualitatively different results.

We now discuss the transitions between the categories of agents in our model. There are three transition probabilities in our model, *p*_1,2_, *p*_2,34_ and *p*_3,4_. These transition probabilities along with the capacity of the public places are the free parameters in our model. When category 1 agents reach their threshold of viral load, a fraction of them transitions into category 2 agents whose viral loads keep on increasing, while the remaining agents remain as category 1, enter the plateau of incubation and finally recover through an exponential decay of their viral load, as shown in Fig. 1. This transition from category 1 to 2 occurs according to the probability *p*_1,2_. The second probability *p*_2,34_ separates the category 2 agents from category 3 and 4 agents, when category 2 agents reach the higher threshold of viral load in Fig. 1. Category 3 and 4 agents are more vulnerable and their viral loads increase after the incubation period and they are admitted to ICU, while category 2 agents recover. The third probability *p*_3,4_ separates category 3 and 4 agents after they are admitted to ICU. There is an initial increase in viral load for both categories, but category 4 agents recover after that, whereas category 3 agents progress towards death.

This model captures a realistic scenario of spread of a pandemic in which infection is transmitted through air and different surfaces. Hence public places are the primary source of spreading the infection. There are several sources of stochasticity in our model, the agents who visit the public places are chosen randomly, and category 1 agents may be chosen repeatedly. The incubation time of the virus in an agent is random within a range, and the probabilities with which an agent transitions from one category to another are also sources of stochasticity. We will analyse the results of our simulation in terms of incremental hospitalization and admission to ICU, comparing with such incremental data released by the health authorities. It will be clear that these three probabilities and the capacity of public places (the four free parameters) are sufficient to match reliable data for the COVID-19 epidemic. We are unaware of any model that is accurate enough to match this kind of hospitalization data.

We now discuss the parameters in the model. They can be classified as *sensitive* and *robust*. The probabilities *p*_1,2_, *p*_2,34_ and *p*_3,4_ are sensitive parameters. Any change of one of these probabilities influences the next probability drastically. For example, if we increase the probability of transition from category 1 to 2, i.e., *p*_1,2_, there will be more category 2 agents in the population and consequently there will be more category 3 and 4 agents. Similarly, if we increase the probability of transition from category 2 to categories 3, 4, i.e, *p*_2,34_, the populations of category 3 and 4 agents increase. The sensitivity of these probabilities results in higher levels of hospitalization and admission to ICU, the two specific data that we use for validating our model. However, one robust aspect of our model is that these probabilities can be tuned by using the hospitalization and ICU data for an initial and relatively short period, and then the trend predicted by the model matches with the actual trend of hospitalization and ICU later.

The robust parameters are related to the different viral load thresholds and the stable levels of viral loads after the thresholds are reached, i.e., the plateau regions in Fig. 1. We denote the viral load threshold for category 1 agents as *t*_1_, that of category 2 agents as *t*_2_, and the threshold for separating category 3 agents from category 4 agents as *t*_3_. Recall that the plateau region for a stable viral load for all agents is chosen as between 3 and 6 days, randomly for each agent. This plateau region is denoted by *P*. We show in the results section that our simulation results do not have much qualitative changes if we vary the parameters *t*_1_, *t*_2_, *t*_3_ and *P*. We summarize these parameters in the following table. The first five parameters are sensitive and the next four are relatively robust.

|  |  |
| --- | --- |
| $p_{1,2}$ | transition probability for hospitalization |
| $p_{2,3,4}$ | transition probability between category 2 and 3, 4 |
| $p_{3,4}$ | transition probability between category 3 and 4 |
| seed | initial seeding of infected agents |
| capacity | capacity of public places |
| $t_1$ | viral load threshold for category 1 agents |
| $t_2$ | viral load threshold for category 2 agents |
| $t_3$ | viral load threshold for category 4 agents |
| $P$ | duration of the plateau or stable viral load |

The model was simulated with ANSI-C multi-threaded code using the OpenMP library. A distributed memory version was tested using the MPI library, however the communication overhead caused lower performance. The simulations were run on the Pawsey supercomputer facility in Perth, Western Australia on two different architectures. The first one is Magnus, a Cray XC40 supercomputer, each compute node includes two Intel Xeon E52690-V3 (Haswell) processors, each with 12 cores, 24 cores in total. The second architecture is Zeus, each node containing two Intel Xeon E5-2680 V4 (Broadwell) 14-core CPUs, for a total of 28 cores. It was necessary to run the longer simulations on Zeus due to a 24-hour time limit for each job on Magnus. The simulation time for a population of 8 millions is about 42 hours, and that for a population of 1.2 millions is about 6 hours. Hence it was difficult to explore the parameter space for the simulation of New York city.

Our simulation strategy is similar to that of Parker and Epstein (7), as we keep track of only the infected agents and not the whole population. The time for each iteration increases as a result as the simulation progresses. Still the efficiency was higher with this approach particularly for larger populations. The memory requirements were also within the 128GB per compute node of both Magnus and Zeus.

## Results

We first show the simulation results for New York City with a population of about 8 million. The hospitalization probability in these simulations is 0.2, i.e., an infected agent transitions to severe with this probability (category 2 agents in our model). Among the agents whose infection levels require hospitalization, the transition to ICU occurs with a probability 0.3 (category 3 and 4 agents in our simulation). And among those who are admitted to ICU, recovery occurs with a probability of 0.5 (category 4 agents). The New York State Government data is shown in Fig 2 and our simulation results are shown in Fig. 3.

**Fig. 2.**
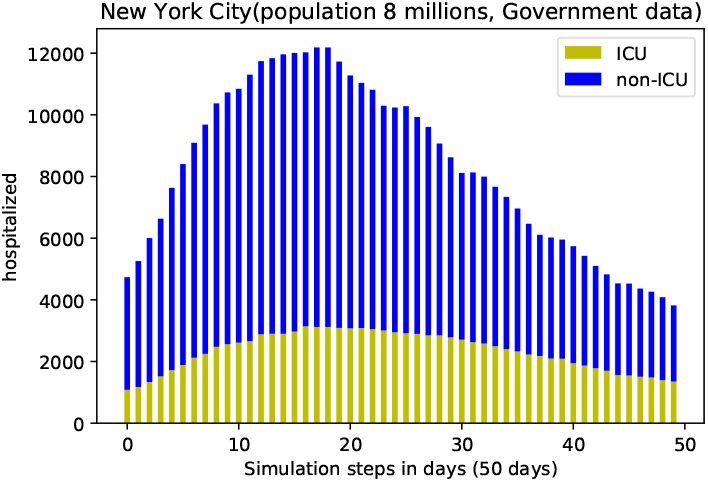
Daily hospitalization and ICU admission summary from NY State Government (15)

**Fig. 3.**
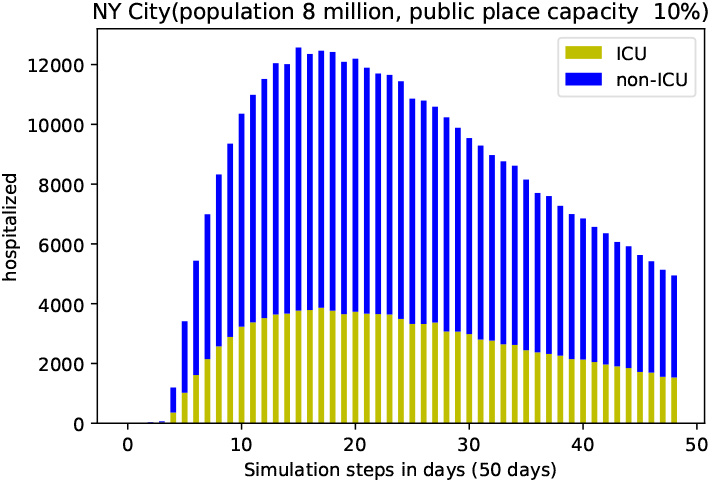
The hospitalization and ICU figures from our simulation.

We also show a direct comparison between government data and simulation data for total hospitalization for NY City in Fig. 4. We show the comparison between government data and simulation data for admission to ICU in Fig. 5. As can be seen our simulation results match quite accurately with the actual data in Fig. 4, except for a steeper rise in simulation results. However, our results for admission to ICU are over-estimates of the actual data. This is due to the fact that we could not explore the parameter space for such a large population due to the very long simulation times. In particular there is a subtle interplay between the sensitive parameters *p*_1,2_ and *p*_2,34_ that we were unable to explore systematically due to very long simulation times.

**Fig. 4.**
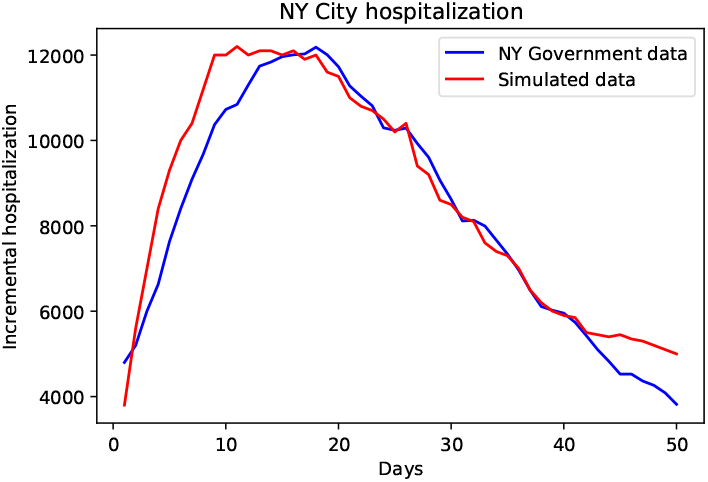
Comparison of government hospitalization data and our simulation results

**Fig. 5.**
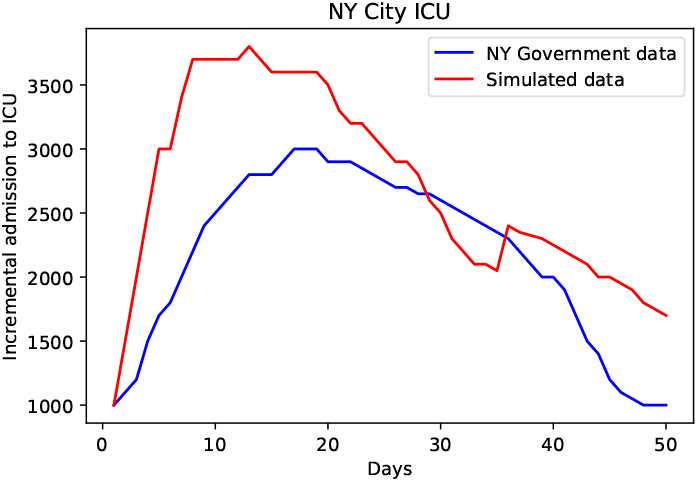
Comparison of government hospitalization data and our simulation results.

The results for Mid-Hudson valley are shown in Figs. 6, 7, 8, and 9. Though the population of Mid-Hudson valley is about 1.35 million, the dynamics of the spread of the infection seems to be very similar to that of NY city, as all the probabilities as well as the public place capacities are exactly the same in these simulations.

**Fig. 6.**
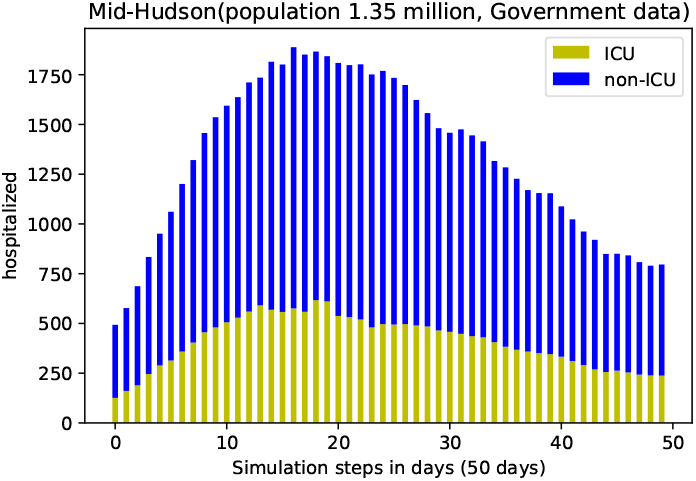
Government hospitalization and ICU data for Mid-Hudson valley

**Fig. 7.**
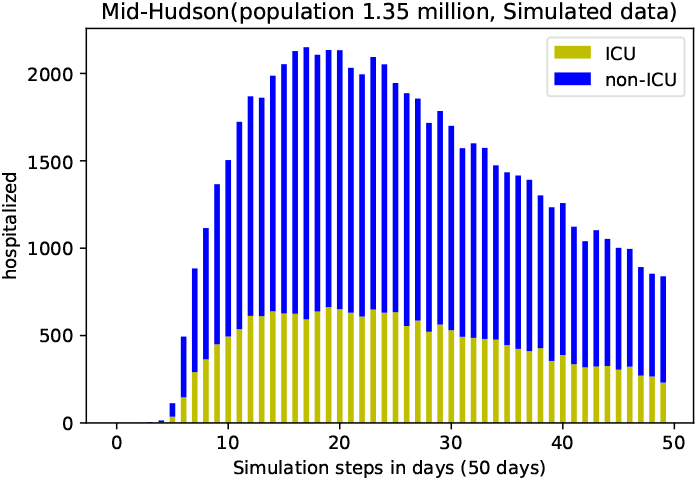
Simulated data for Mid-Hudson valley.

**Fig. 8.**
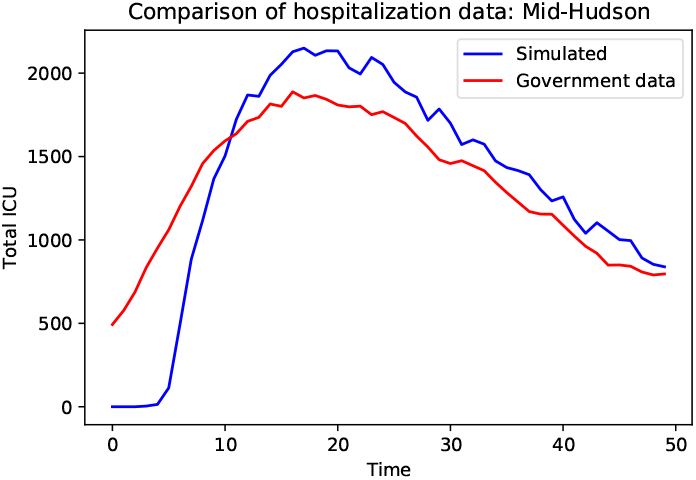
Comparison of government data and simulated data for hospitalization for Mid-Hudson valley.

**Fig. 9.**
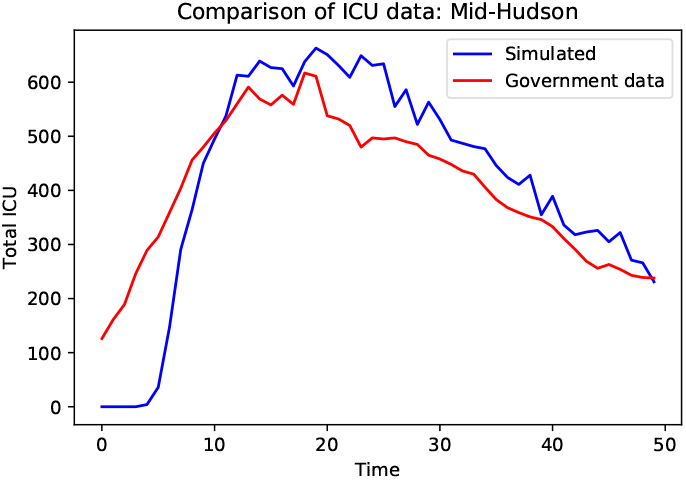
Comparison of government data and simulated data for admission to ICU for Mid-Hudson valley.

Next, we show the simulation results for a smaller population and compare it with the government data for hospitalization and admission to ICU. This simulation is for the Western New York region with a population of 1.8 million. The overall hospitalization and admission to ICU are much smaller in number compared to that of Mid-Hudson valley, which has a comparable population. These results for the WNY region are shown in Figs. 10, 11, 12 and 13.

**Fig. 10.**
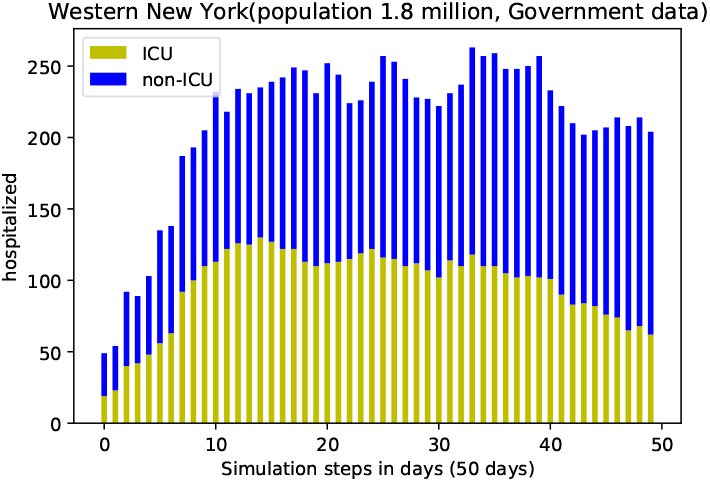
NY Government hospitalization data for Western NY.

**Fig. 11.**
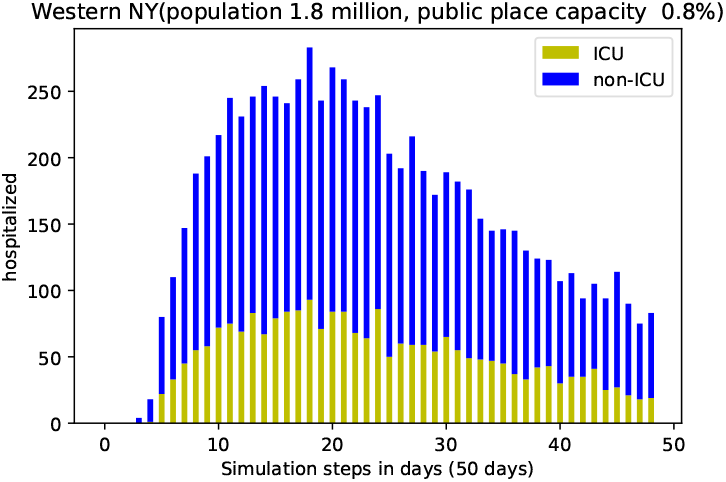
Simulated data for Western NY region.

**Fig. 12.**
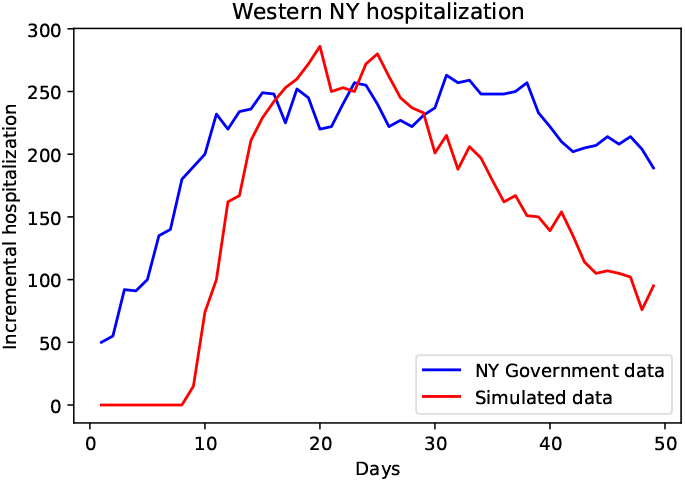
A comparison of simulated data and NY Government hospitalization data for the Western NY region.

**Fig. 13.**
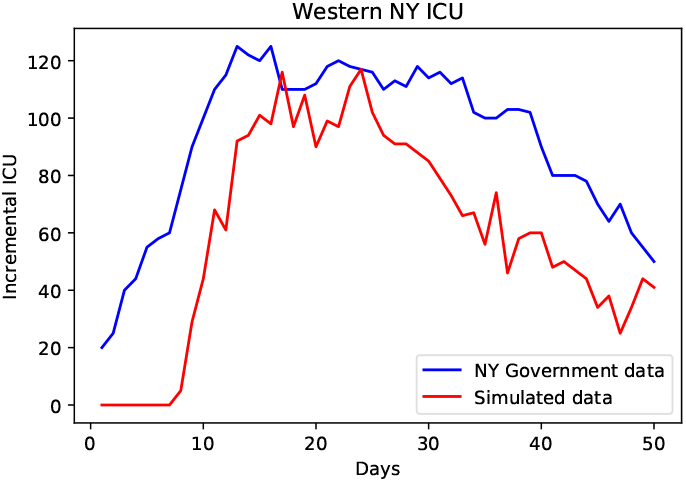
A comparison of simulated data and NY Government ICU data for the Western NY region.

We summarize the simulation probabilities and public place capacities for NY City, Mid-Hudson valley and Western New York region in the following table. Public place capacities are shown as a percentage of the population. All simulations were run with initially 0.01% of infected agents (seed infection).

| Region | $p_{1,2}$ | $p_{2,3,4}$ | $p_{3,4}$ | Public place |
| --- | --- | --- | --- | --- |
| NY City | 0.2 | 0.3 | 0.5 | 10% |
| Mid-Hudson | 0.2 | 0.3 | 0.5 | 10% |
| Western NY | 0.017 | 0.35 | 0.5 | 1% |

We next explore the effects of varying the robust parameters in our model with respect to the simulation of the Western New York region. Figs. 14 and 15 compare the simulation results when the total capacity of public places is 1% and 5% respectively. The changes in the simulation results are not significant. This is due to the fact that though a larger capacity of public places potentially exposes more agents to infection, we have assumed that the initial viral loads are small and hence the increase in viral load over time is more important. In other words, even with a smaller capacity in public places, the number of agents that can be accommodated in public places does not change significantly as the duration of stay of an agent in a public place is only about 10 minutes according to our calibration.

**Fig. 14.**
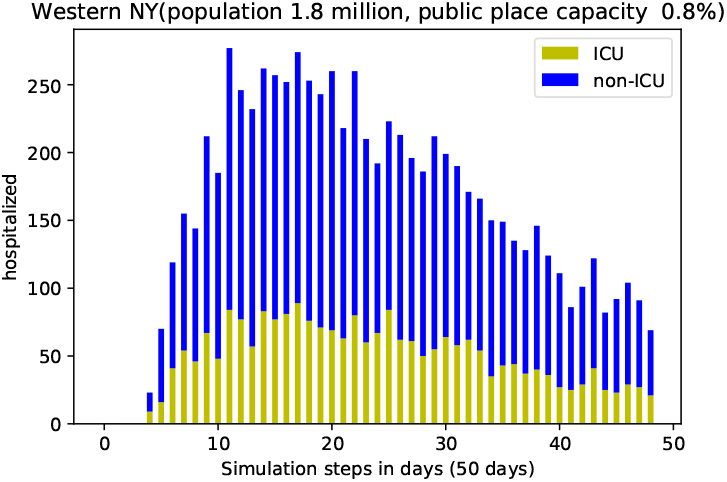
Simulation for 50 days with public place capacity 1% of the population.

**Fig. 15.**
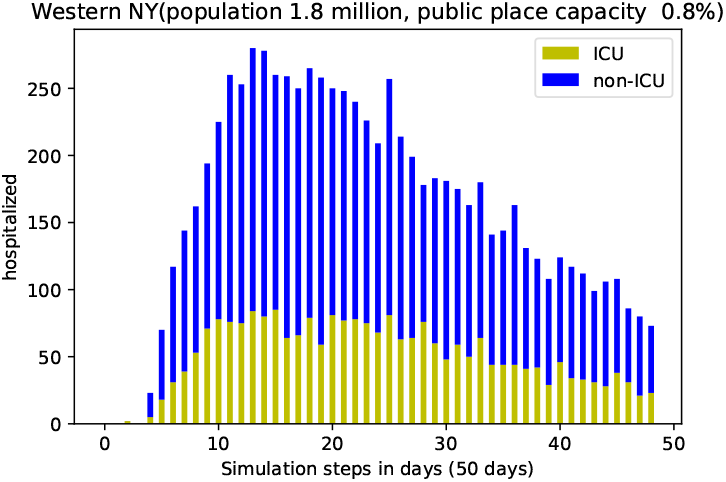
Simulation for 50 days with public place capacity 5% of the population.

We have shown simulation results by doubling the transition thresholds between different categories of agents in Fig. 16. We also vary the incubation period of the virus randomly between 6-12 days in Fig. 17. Again, there are no significant changes, except for shifting the hospitalization and ICU admission rates.

**Fig. 16.**
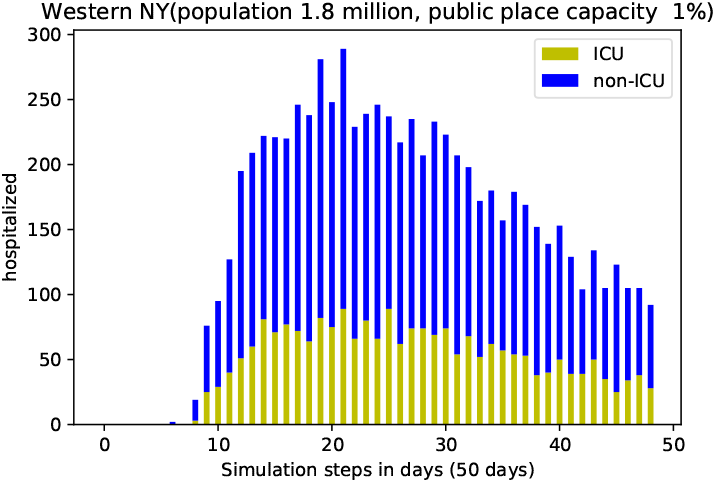
Simulation for 50 days with public place capacity 1% of the population. The viral load thresholds are double of those compared to Fig 11,Fig 14 and Fig 15

**Fig. 17.**
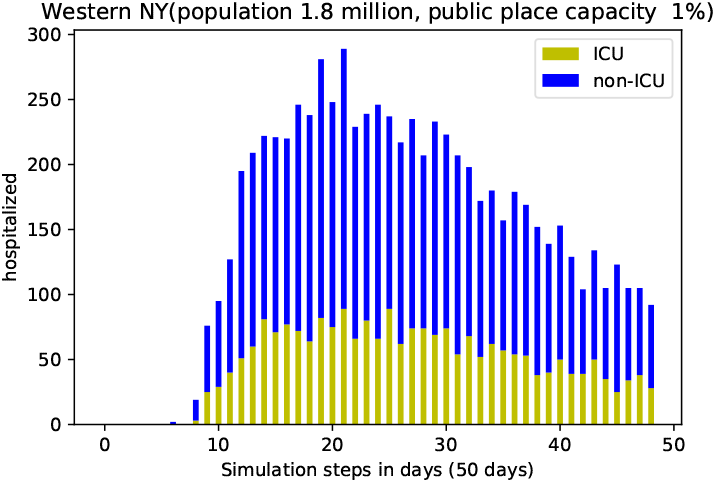
Simulation for 50 days with public place capacity 1% of the population. The viral incubation period is 6 − 12 days.

It is possible to study lockdowns easily in our model. We can choose to prevent the agents from visiting public places for a certain number of simulation steps and that has the effect of lockdowns. For example Fig. 18 shows a simulation for Western New York, with lockdown imposed between days 8-16. As a result, the incremental hospitalization and ICU admission rates drop to zero after a few days, as the early lockdown has not increased the infection levels of infected agents to levels of category 2,3 or 4 agents. On the other hand, a later lockdown would reduce the hospitalization and ICU admission rates.

**Fig. 18.**
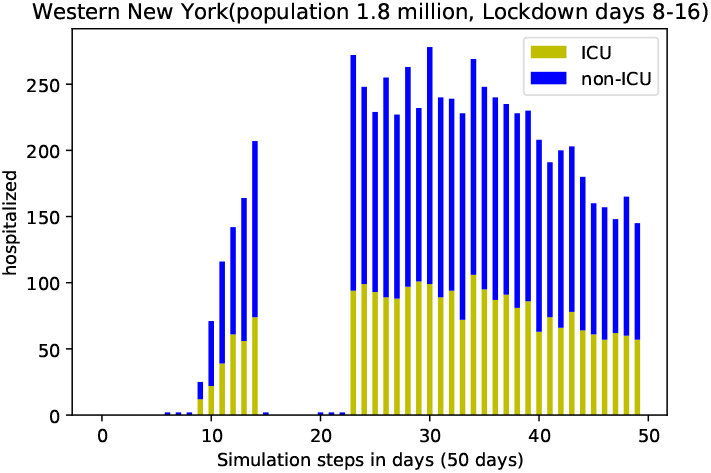
Simulation for Western New York, with lockdown imposed between days 8-16

## Conclusions

We have presented a simple agent-based simulation model in this paper that can match the actual hospitalization and ICU admittance data quite accurately. We would like to highlight that this simple model can be enhanced in several different ways to reflect the actual social interactions and hence the spread of a pandemic in the real world. Though this model is inspired by the works by Epstein (6) and Parker and Epstein (7), their model is of global scale, whereas our model is targeted for much smaller communities in cities and regions. The possible enhancements of our model can motivate the study of spread of a pandemic more precisely. For example, it is possible to impose a spatial grid on our home communities and public places to reflect the different parts of a city or community. It is even possible to simulate movable public places like public transportation and subway systems. A spatial grid will also enable imposing selective lockdown measures on different locations in the community, and study the effect of social distancing more precisely. As the results for NY City with a large population and Mid-Hudson with a much smaller population show, perhaps the location of Mid-Husdon is the reason for similar spread of the infection in these two communities. On the other hand though Western New York has a similar population as Mid-Hudson, the dynamics of spread of infection is quite different there, as is evident from both the government data and our simulation results. These kind of model findings can help suggest directions for further investigations of the pandemic spread in Western New York and other areas.

We have assumed that an agent stays at a public place only for a short time, in other words our assumption is that the public places are homogeneous. It is also possible to group public places in our model according to shorter and longer stay, e.g., grocery stores, public transportation and offices. That will again facilitate a finer scale study of the spread of the pandemic. Our model has the capability to make forward predictions on the access needed to public health facilities, and that can help the local and state governments to prepare for the spread of the pandemic. The fine tuning of the parameters by noting the initial spread of the pandemic will help in predicting the medium term requirements of the public health facilities.

## Data Availability

All data and computer code used in this paper are available on request. As such all data in this paper are generated using computer simulation, so there is no data repository.

## Notes

### Competing Interest Statement

The authors have declared no competing interest.

### Funding Statement

There is no direct funding, we have done this work as academics in our research time.

### Author Declarations

This research is purely computer simulation using publicly available data and hence there was no need for approval or exemption.

